# Extract, Transform, Load Framework for the Conversion of Health Databases to OMOP

**DOI:** 10.1101/2021.04.08.21255178

**Authors:** Juan C. Quiroz, Tim Chard, Zhisheng Sa, Angus Ritchie, Louisa Jorm, Blanca Gallego

**Author notes:** Corresponding author: Juan C. Quiroz, Centre for Big Data Research in Health, Level 2, AGSM Building, G27, Botany St, Kensington NSW 2052.

## Abstract

**Objective:** Develop an extract, transform, load (ETL) framework for the conversion of health databases to the Observational Medical Outcomes Partnership Common Data Model (OMOP CDM) that supports transparency of the mapping process, readability, refactoring, and maintainability.

**Materials and Methods:** We propose an ETL framework that is metadata-driven and generic across source datasets. The ETL framework reads mapping logic for OMOP tables from YAML files, which organize SQL snippets in key-value pairs that define the extract and transform logic to populate OMOP columns.

**Results:** We developed a data manipulation language (DML) for writing the mapping logic from health datasets to OMOP, which defines mapping operations on a column-by-column basis. A core ETL pipeline converts the DML in YAML files and generates an ETL script. We provide access to our ETL framework via a web application, allowing users to upload and edit YAML files and obtain an ETL SQL script that can be used in development environments.

**Discussion:** The structure of the DML and the mapping operations defined in column-by-column operations maximizes readability, refactoring, and maintainability, while minimizing technical debt, and standardizes the writing of ETL operations for mapping to OMOP. Our web application allows institutions and teams to reuse the ETL pipeline by writing their own rules using our DML.

**Conclusion:** The research community needs tools that reduce the cost and time effort needed to map datasets to OMOP. These tools must support transparency of the mapping process for mapping efforts to be reused by different institutions.

## INTRODUCTION

Electronic health records (EHRs), administrative data, clinical registries, and linked data enable observational studies and evidence-based research that leverage the data of large and heterogenous populations [1–3]. The growing availability of EHR-linked biobanks also facilitate research and implementation of patient phenotyping and personalized medicine [4]. Depending on the location, context, and purpose of the data, different datasets store information using different structure and semantics, which makes conducting analysis across them a challenge. Common data models offer a solution by standardizing the structure and semantics of data. The Observational Medical Outcomes Partnership Common Data Model (OMOP CDM), managed by the Observational Health Data Sciences and Informatics (OHDSI), continues to be one of the leading common data models for leveraging clinical and administrative health data for research purposes [5]. Since its introduction, health databases have been mapped to OMOP CDM, including EHRs [6–10], claims datasets [9,11,12], biospecimen data [13], and registries [14].

Converting routinely collected health data to OMOP is driven by the aims of: (1) efficiency–reuse of analytics tools and software, (2) transparency and reproducibility—compare findings from different data sources with a variety of methods without sharing the actual data and protecting patient privacy, and (3) scalability—conducting studies by leveraging the data from multiple locations and settings [5,15]. The need for OMOP also stems from health databases originating or being designed to track patients within a hospital, mainly for administrative purposes such as billing and managing claims, not for conducting observational studies or other study designs [2,16]. At its best, data in OMOP allows for multicenter observational studies, allowing for models to be externally validated across health datasets over the world [5,17,18].

Converting a dataset to the OMOP CDM entails the development of an extract, transform, load process (ETL), which converts both the structure and semantics of the original data to the standards defined by the OMOP CDM. Conceptually, the mapping process identifies how fields in the source health datasets relate to the fields in OMOP CDM and the data transformations that need to take place. Resources currently available to a team or institution interested in converting their data to OMOP include: OMOP experts, other users who have mapped data to OMOP, OHDSI web forums, and private companies that perform the ETL process for a fee.

The OHDSI research community has developed tools for supporting the mapping process, such as a tool for understanding the source database (White Rabbit) and a tool for documenting the mapping requirements (Rabbit in a Hat) [19]. However, the graphical approach of Rabbit in a Hat does not scale to a large number of columns and cannot deal with complex mapping logic involving multiple source tables—commonly encountered in the mapping of sophisticated, proprietary relational databases used by commercial EHRs. Tools are required that improve the ETL process, enable mapping efforts from research institutions to be shared, and standardize the writing of mapping operations for simple and complex datasets.

We propose an ETL framework for mapping health datasets to OMOP CDM that is driven by a data manipulation language (DML) that organizes SQL in YAML, a widely used human-readable serialization language. The DML enforces a column-by-column definition of data manipulation. An ETL pipeline, accessible via a web application, converts the rules written with the DML into an ETL SQL script that can be executed in development environments. This approach improves the readability of mapping rules, provides flexibility in the process, allows the mapping rules to serve as documentation and as input to the ETL framework, and standardizes the writing of ETL operations allowing for sharing and curation of mapping logic. The ETL framework was developed as part of our ongoing work mapping Cerner Millennium (Cerner Corporation, Kansas City, MO, USA, “CERNER”) electronic health records used by Australian Local Health Districts to OMOP CDM, and all examples presented in this paper use this CERNER to OMOP conversion to showcase the DML.

## EXTRACT, TRANSFORM, LOAD FRAMEWORK

Figure 1 illustrates the architecture of our ETL process from a source database to the target OMOP CDM dataset. A compiler reads rules written in our data manipulation language (DML) and generates an ETL SQL script containing all the executable operations to extract, transform, and load the data from the source database to OMOP. The ETL script can then be used in any development environment. Access to the source code of the compiler is available in https://github.com/clinical-ai/omop-etl.

**Figure 1.**
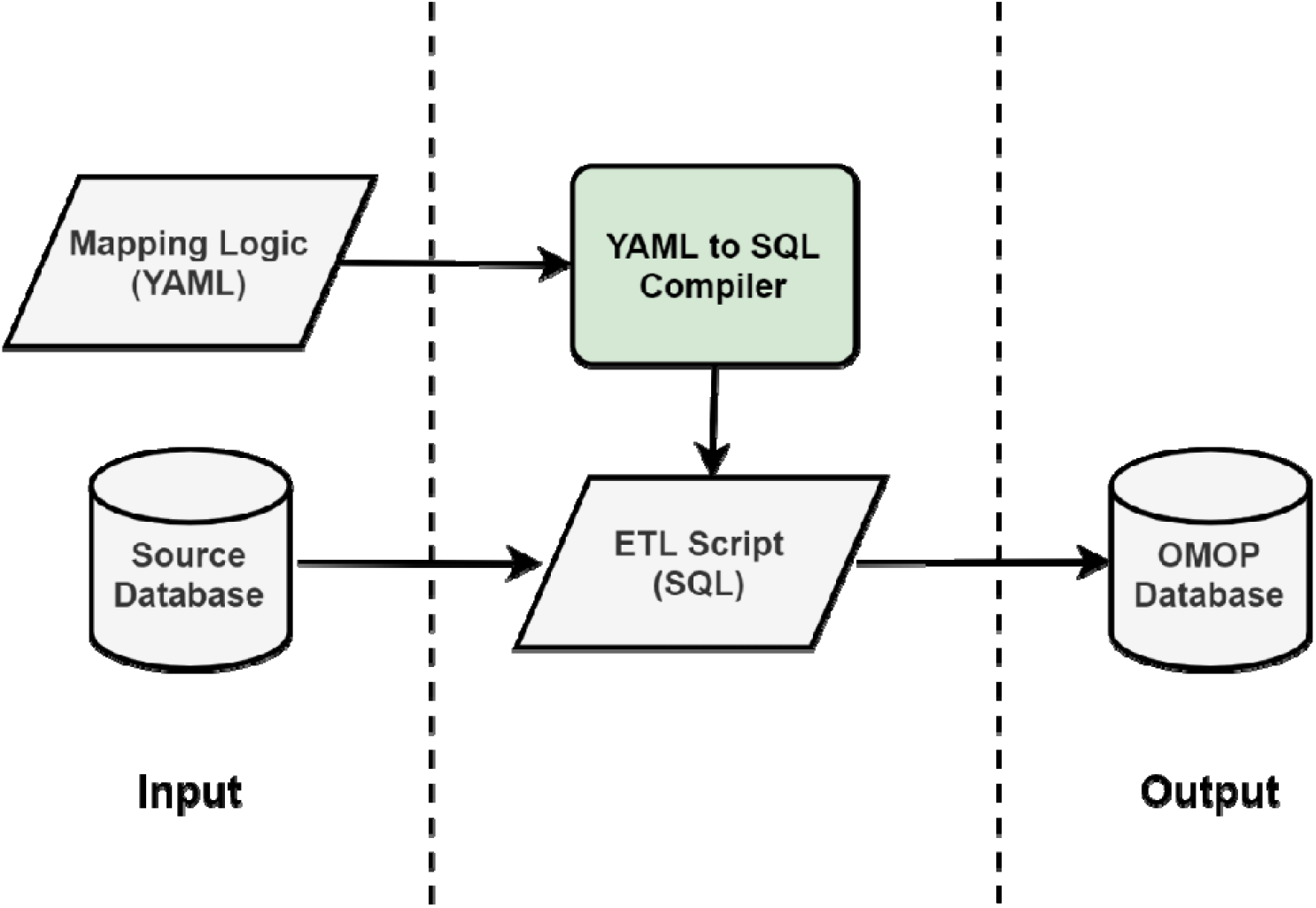
Extract, transform, load (ETL) framework that is driven by a new domain manipulation language (DML) that uses YAML and PostgreSQL syntax. A compiler generates an ETL script from the mapping operations defined in YAML. The ETL script contains all the executable operations to map from the source database to OMOP.

### Data Manipulation Language

The DML uses YAML and PostgreSQL syntax. YAML is a human-readable data format, commonly used for storing configuration settings and metadata of software programs. The DML uses YAML key-value pairs to define the source data, the target OMOP tables and columns, and the extract, transform, and load operations to map from source data to OMOP. Information about the source schema does not need to be explicitly provided, as the rules written with the DML contain all the information needed to extract data from the source dataset.

YAML was chosen because it is easy to read and flexible to handle complex relationships. The definition of a YAML schema in our DML also provides validation of YAML files, ensuring that rules written by users with our DML follow the right structure, standardizing the writing of OMOP mapping rules. The use of PostgreSQL syntax allows users to use all functions and operations supported by PostgreSQL, without forcing users to learn a new SQL language.

The source-to-OMOP ETL operations are organized by OMOP table (i.e. PERSON, OBSERVATION_PERIOD, DRUG_EXPOSURE). Each YAML file describes the mapping logic for a target OMOP table (Figure 2) and contains three sections: (1) name of the OMOP table being mapped (YAML field *name*), (2) definition of primary keys used by the ETL framework to manage the load (insert) operations (YAML field *primary_key)*, and (3) mapping rules for each column in the targeted OMOP table (YAML field *columns*).

**Figure 2.**
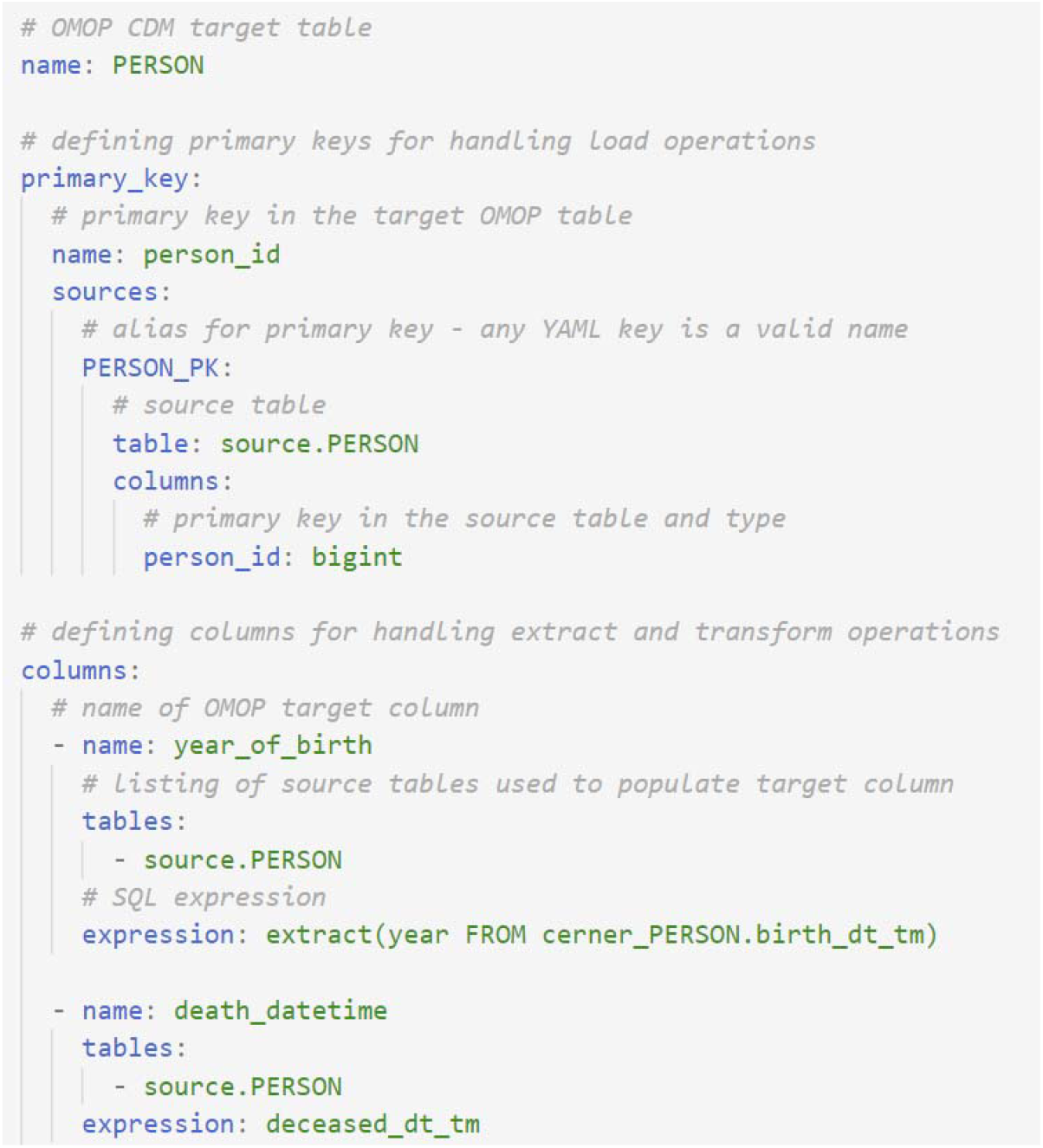
Rules for mapping from the CERNER PERSON table to the OMOP PERSON table, with rules defined for two columns of the OMOP PERSON table: year_of_birth and death_datetime. For each target table, the mapping rules are defined on a column-by-column basis using SQL snippets organized into YAML fields. Mapping a table requires three sections: (1) name of the OMOP table being mapped, (2) definition of primary keys used by the ETL framework to manage the load (insert) operations, and (3) mapping rules for columns in the OMOP table.

### Defining Primary Keys

OMOP tables are populated with the data from one or more source tables. For simple datasets, there may be a one-to-one relationship between source and OMOP tables. However, in the case of most datasets (such as EHRs), fields in OMOP tables will likely be populated with the data from multiple source tables. In cases where an OMOP table is populated with the data from a single table (as in the case of populating the OMOP Person table from the CERNER Person table), we derive the primary key for the OMOP table from the primary key in the source table. This is under the *primary_key* YAML field (Figures 2 and 3). If the primary key of the source table is not compatible with the type of the destination field, then our ETL framework automatically handles an appropriate conversion.

**Figure 3.**
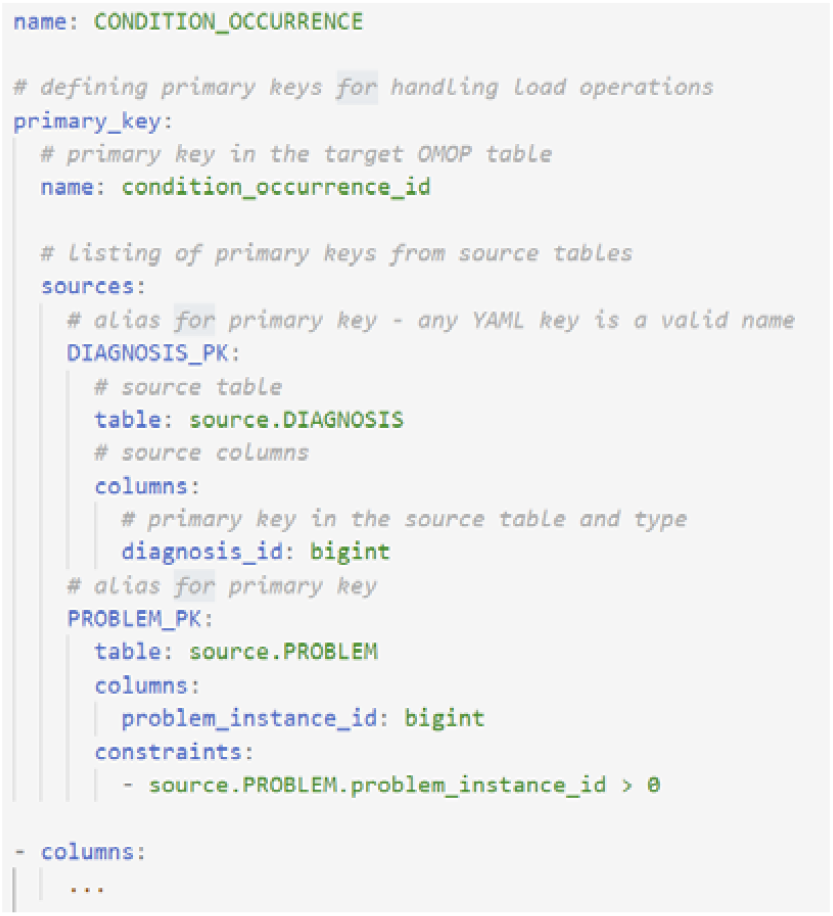
Multiple primary keys can be defined under the primary key field. The primary keys are used to handle the load operations. Multiple primary keys are necessary when the data from multiple source tables are used to populate a target OMOP table.

In cases where the data from multiple source tables are used to create new records—i.e. the OMOP table represents a union of two or more source tables—multiple primary keys can be defined (Figure 3). The defined primary key can then be associated with the mapping logic of a column, ensuring that the column gets inserted using the associated primary key.

### Defining Columns

The information needed to define the extract and transform operations from source data to an OMOP column are: (1) the name of the targeted OMOP column (YAML field *name*), (2) a listing of one or more source tables containing the data needed to populate the target field (YAML field *tables*), and (3) an SQL expression defining how one or more fields from the source table(s) map to the OMOP field (YAML field *expression*). Figure 2 shows the extract and transform operations to populate two columns (year_of_birth and death_datetime) of the PERSON OMOP table. The *expression* field supports PostgreSQL syntax, enabling the use of all functions and syntax supported by PostgreSQL (see Figure 4). For complex mapping logic, the *tables* field also supports SQL select queries.

**Figure 4.**
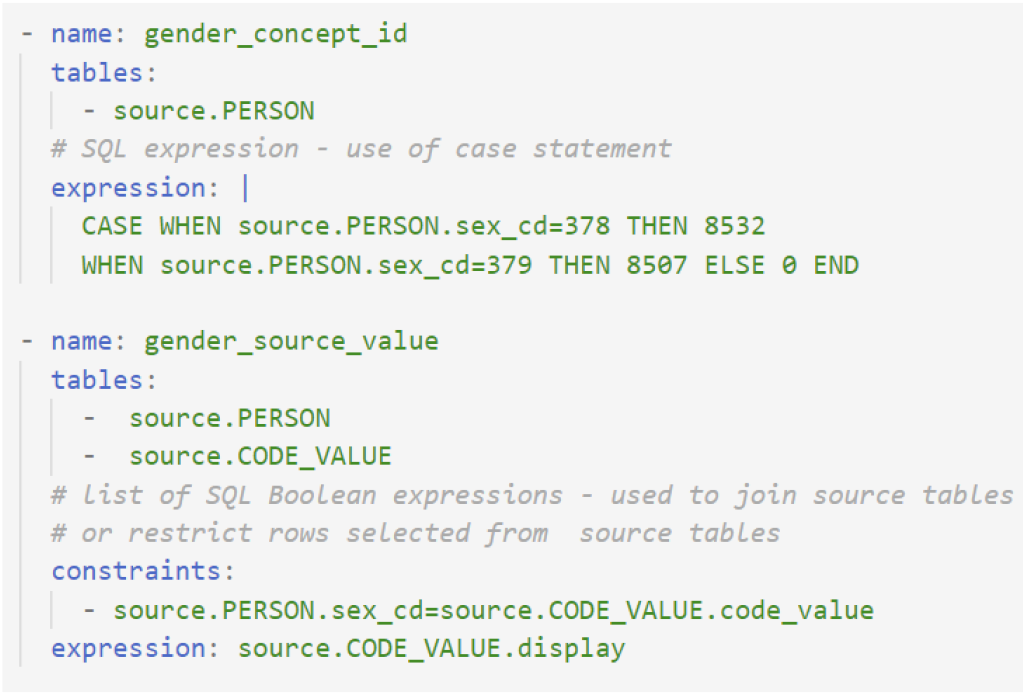
Multiple primary keys can be defined under the primary key field. The primary keys are used to handle the load operations. Multiple primary keys are necessary when the data from multiple source tables are used to populate a target OMOP table.

When data from multiple source tables is needed to populate an OMOP field, as illustrated in Figure 4, the *constraints* field (a Boolean PostgreSQL expression) can be used to define how the source tables are joined. The constraint field can also be used to filter rows from the source tables that meet the conditions listed under *constraints*. Constraints can also be used with a single source table and to restrict fields on the basis of primary keys (as shown in Figure 3).

### Multiple Rules Per Column

The mapping logic defines operations on a column-by-column basis, but in complex cases, all the rows needing to be mapped may be updated using separate rules. This is especially useful for breaking down complex logic into rules that map subsets of a single column. Figure 5 shows an example of two rules mapping source data to the condition_start_date OMOP column in the CONDITION_OCCURRENCE OMOP table. In the example, one rule is used to map diagnosis events and the second rule maps problem events to OMOP condition occurrence events. When multiple primary keys are defined, the alias of the primary key is used to indicate the primary key corresponding to a particular rule.

**Figure 5.**
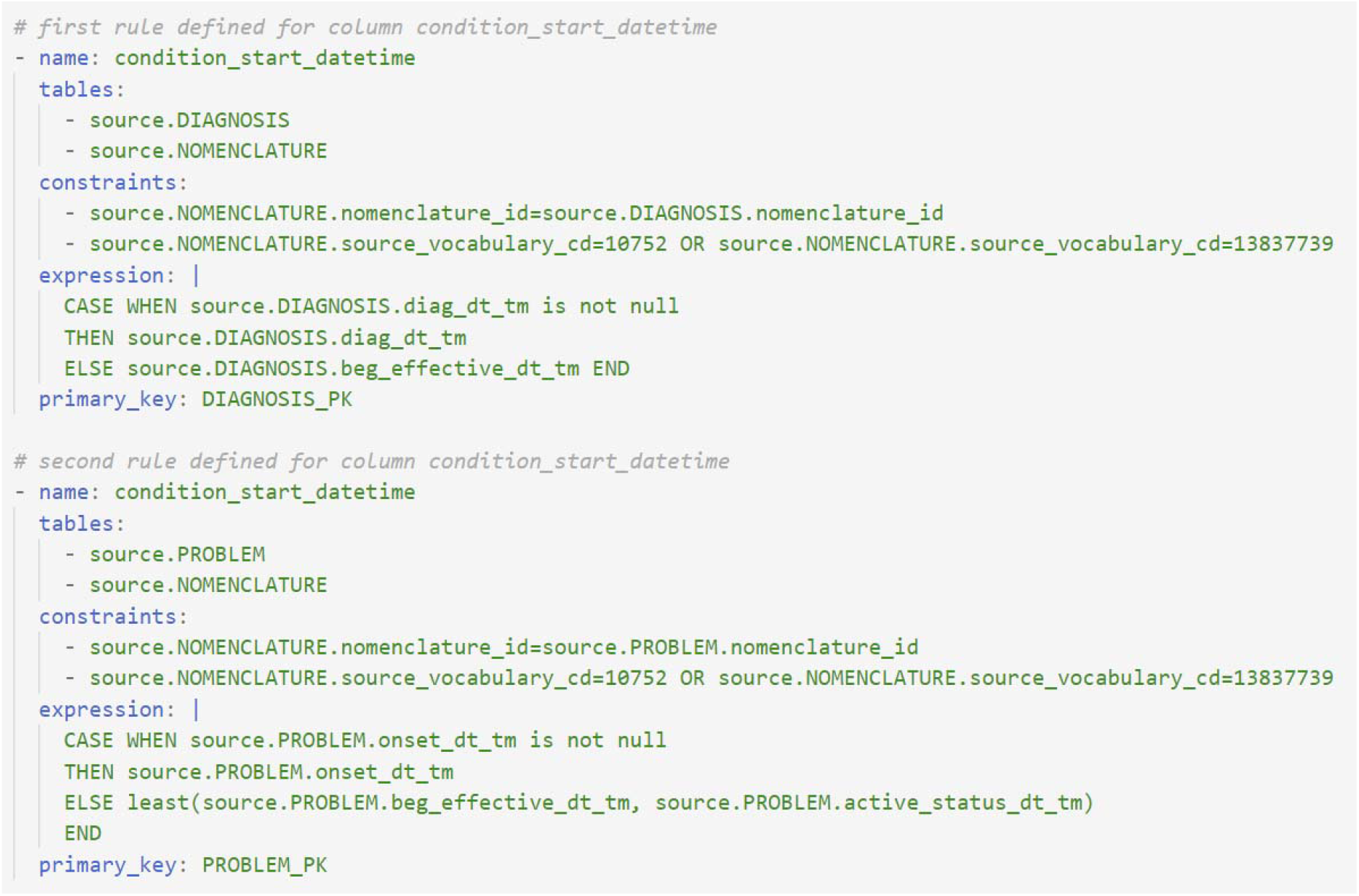
Mapping rules illustrating complex logic relying on multiple source tables to populate the OMOP CONDITION_OCCURRENCE table. Multiple rules can be written to map source data to a single OMOP column, dividing complex logic into queries that are easier to read and debug.

### Reducing Repetition in Mapping Logic

The use of YAML allows mapping rules to be written using YAML anchors and aliases. Anchors and aliases enable YAML fields to be defined once and reused multiple times, removing repetition in the YAML files and resulting in ETL operations that are easier to read in comparison to SQL. Figure 6 shows an example of an anchor being defined (“default_values”), which is subsequently used in the two columns year_of_birth and death_datetime.

**Figure 6.**
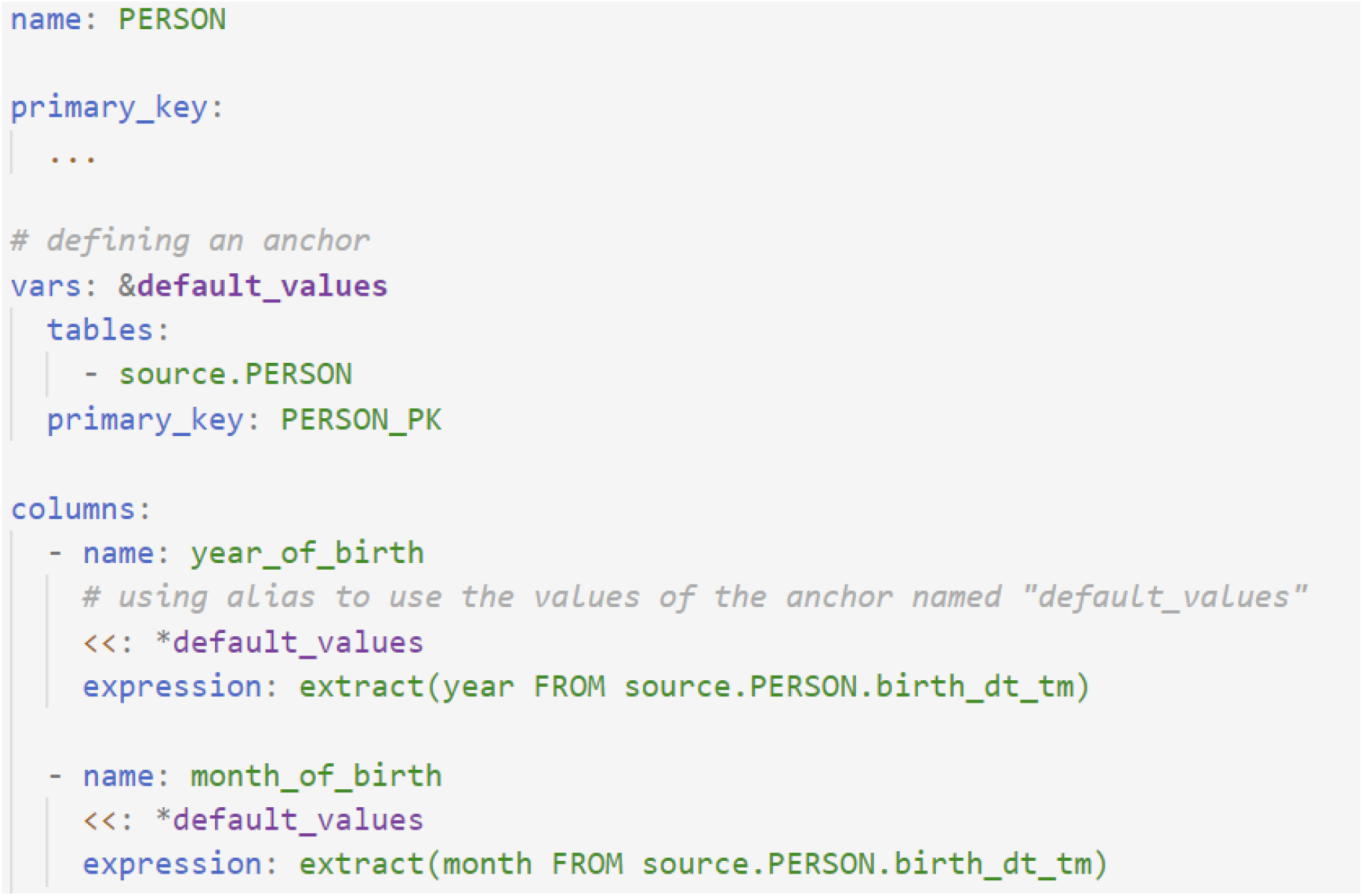
Rules for mapping from the CERNER PERSON table to the OMOP PERSON table, using YAML anchors to define a set of default values to be used by the column fields.

### Web Application

A web application (http://www.omop.link/) provides end-users with API access to the YAMLto-SQL compiler, including an editor for writing mapping logic, validating the DML, and default pre-filled OMOP v6 YAML for OMOP tables (See Figure in Appendix for web editor view). The web application converts the YAML ETL operations to an ETL SQL script containing all the executable operations to extract, transform, and load the data from the source database to OMOP. No information about the source database schema is needed, as all the necessary logic is contained in the YAML content. The use of our DML and our web application thus has the potential to support the mapping from any source SQL database to OMOP. The web application uses React, Bootstrap, and the Monaco Editor for the front-end. The backend uses the Python FastAPI library.

### Validation

Validation of our ETL framework consisted of 14 test cases (Appendix - Table 1) representative of commonly used combinations of features of the DML (for example, cases where the target column is a foreign key or the target table involves primary keys from multiple source tables). Once these test cases were identified, we chose columns in the OMOP CDM tables that were appropriate candidates for each test case. We then manually generated synthetic CERNER source data and the expected mapped OMOP data for the 14 test cases for comparison against the output of the ETL framework (see Figure in Appendix 2 for a diagram of how the CERNER source test data was associated with OMOP tables). Finally, we wrote the corresponding YAML files using our DML and generated an SQL script using our compiler. The test CERNER data was converted to OMOP by executing the SQL script against a PostgreSQL database. The resulting OMOP data was then compared with the manually mapped data.

**Table 1.** Test cases to validate the ETL framework, with the collective set of test cases evaluating all features of the ETL framework.

| Test case | Primary keys from multiple source tables | Column is Primary Key | Column is Foreign Key | Expression | Column involves constraints | OMOP Target Column |
| --- | --- | --- | --- | --- | --- | --- |
| 1 |  | ✓ |  | Simple | ✓ | PERSON<br>person_id |
| 2 |  |  |  | Simple |  | PERSON<br>death_dt_tm |
| 3 |  |  |  | Complex |  | PERSON<br>year_of_birth |
| 4 |  |  |  | Constant |  | PERSON<br>gender_source_concept_id |
| 5 |  |  |  | Simple | ✓ | PERSON<br>gender_source_value |
| 6 |  | ✓ |  | Simple | ✓ | VISIT_OCCURRENCE<br>visit_occurrence_id |
| 7 |  |  | ✓ | Simple | ✓ | VISIT_OCCURRENCE<br>person_id |
| 8 | ✓ | ✓ |  | Simple | ✓ | CONDITION_OCCURRENCE<br>condition_occurrence_id |
| 9 | ✓ | ✓ |  | Simple | ✓ | LOCATION<br>location_id |
| 10 | ✓ |  |  | Simple | ✓ | CONDITION_OCCURRENCE<br>condition_concept_id |
| 11 | ✓ |  |  | Complex |  | LOCATION<br>zip |
| 12 | ✓ |  |  | Constant |  | LOCATION<br>state |
| 13 | ✓ |  |  | Simple | ✓ | CONDITION_OCCURRENCE<br>condition_concept_id |
| 14 | ✓ |  | ✓ | Simple | ✓ | CONDITION_OCCURRENCE<br>person_id |

## DISCUSSION

Our work was driven by the need for tools that support ETL processes when mapping health datasets to OMOP. This new ETL design approach is driven by the design principles of column-by-column mapping of data to OMOP, maximizing readability, and standardizing the writing of ETL operations for mapping datasets to OMOP. Our framework divides the ETL process into (1) a core ETL pipeline that reads and executes extract-transform operations from YAML files, and (2) YAML files that organize SQL snippets defining ETL operations on a column-by-column basis. This approach makes the core ETL pipeline reusable to other research groups, accessible via a web application, with users being responsible for writing the YAML files with the ETL operations. It also enables our tool to be used with sensitive health datasets, as our web application only relies on the YAML content to generate the ETL script that can be used in various deployment and secure environments.

In contrast to an ETL process written entirely in a programming language such as Python (or otherwise), our approach stores the ETL operations in separate YAML files, making them easily accessible and promoting transparency of the ETL process. In contrast to an ETL process written entirely as an SQL script, the structure of the YAML configuration leads to readable ETL logic by defining operations on a column-by-column basis. Our proposed framework provides an alternative to existing ETL pipelines and can be used in combination with existing ETL frameworks.

The column-by-column processing of our framework tackles the spaghetti query anti-pattern [20], as forcing users to think of data manipulation on a column-by-column basis is a form of divide-and-conquer that encourages users to write simple queries. This is in contrast to mapping data on a row-by-row basis, with a single complex (spaghetti) query defining the transformations for all the columns of a table. Writing rules on a column-by-column basis also allows for mapping logic to be written for a subset of OMOP, with later additions simply consisting of adding column mappings to the YAML files. Hence, logic added to map new columns does not affect or change prior column definitions, making it less likely that bugs will be introduced as a result of revising mapping logic. Finally, the column-centric approach also allows flexibility in the mapping process, afforded by both the SQL and YAML syntax, enabling the same logic to be written in different ways.

The readability of mapping logic written in our DML facilitates debugging, refactoring, and understanding by users other than the original writer of the mapping logic. This serves to minimize the technical debt associated with maintaining the mapping logic and “bad smells” (code suggestive of design problems) of complex SQL queries [21,22]. The readability of our DML also enables the sharing of the mapping logic with stakeholders who are not database experts and across institutions mapping data to OMOP. Sharing mapping logic, or subsets thereof, is as simple as sharing the YAML files.

Our web application and ETL framework have several advantages for health datasets and sensitive data. The web application does not need access to the source data. Users have the freedom to write rules inside a secure environment or within firewalls meant to protect sensitive health data. Because rules do not contain sensitive information, they can be uploaded to the web application, converted into an ETL SQL script, which can then be executed in any number of environments.

An OMOP CDM goal is to increase the transparency and replicability of real-world evidence research. If the first step in performing these research studies—the mapping of the source data to OMOP—is not transparent, reproducibility may be compromised. Our ETL framework and DML prioritize mapping logic standardization and readability, promoting transparency and reproducibility.

### Comparison With Existing Literature

Most publications reporting on the mapping of specific health datasets to OMOP, simply document the data they map, challenges encountered, and data validation of the mapped data [6–11,23]. Some studies report replications of studies comparing results with the source data and the data mapped to OMOP (to identify biases or discrepancies introduced as part of the translational process) [24]. However, these are of limited value to health institutions or research groups attempting to map third datasets to OMOP.

Rabbit in a Hat is a graphical documentation tool for generating mapping requirements [19]. Mappings are defined by drawing arrows from source tables to the corresponding columns in OMOP tables. The problem with this approach is that it is not scalable for a large number of columns and it does not lend itself to complex mapping logic (multiple source tables used to populate one OMOP column). As such, it is only a useful tool in documenting the mapping of simple datasets where the majority of the mappings are one-to-one from source column to OMOP column. In addition, the result is a mapping document (a requirements specification), that must then be coded by software engineers or a database expert.

### Limitations

For complex health databases, such as CERNER EHR, the most challenging aspect of mapping data to OMOP is determining the logic for changing the semantic meaning of data to fit within the OMOP CDM constraints. Our ETL framework does not address this need, and instead focuses on the standardization of mapping logic and improving transparency of the ETL process. However, the research community and health institutions need more tools to support this process, such as OHDSI’s White Rabbit.

Vocabulary mapping is a particularly challenging aspect that is commonly highlighted in studies mapping to OMOP or standardizing medical terminologies [9,25]. As such, improvements to tools such as Usagi [26] and natural language processing approaches are needed to support vocabulary mapping. This effort has been left for future work. The Web API tool currently generates an ETL PostgreSQL script. However, this can be easily extended to generate scripts for other relational database management systems (MySQL, Oracle) and in more dynamic scripting languages (Python, R) as future work.

## CONCLUSION

The ETL framework proposed in this study was developed as part of our effort to map the electronic health records of local health districts in Australia to OMOP. Our team is developing new tools to improve the current mapping efforts to OMOP, enabling institutions to map datasets to OMOP at a lower cost and complexity, helping to build capacity for health services to partner in advancing data science. The design of the ETL framework is also driven by the goals of the OHDSI community, to increase transparency and reproducibility of research, and the sharing of tools that will facilitate cross-institutional research. Our ETL framework achieves this through a DML that is readable and easy to share, and a web application enabling research teams to use our DML for mapping their data to OMOP.

## Data Availability

NA

## Author contributions

JCQ, TC, ZS, and BG contributed to the conception and design of the framework. TC did the development of the framework and execution of the validation. BG manually mapped data for validation. JCQ drafted the initial manuscript. All authors contributed to critical revisions of the manuscript. All authors approved the final draft.

## Acknowledgements

Dr. Malcolm Gillies and Dr. Oscar Perez-Concha for feedback on ETL processes.

## APPENDIX

### ETL Framework Web Application

**Figure 1.**
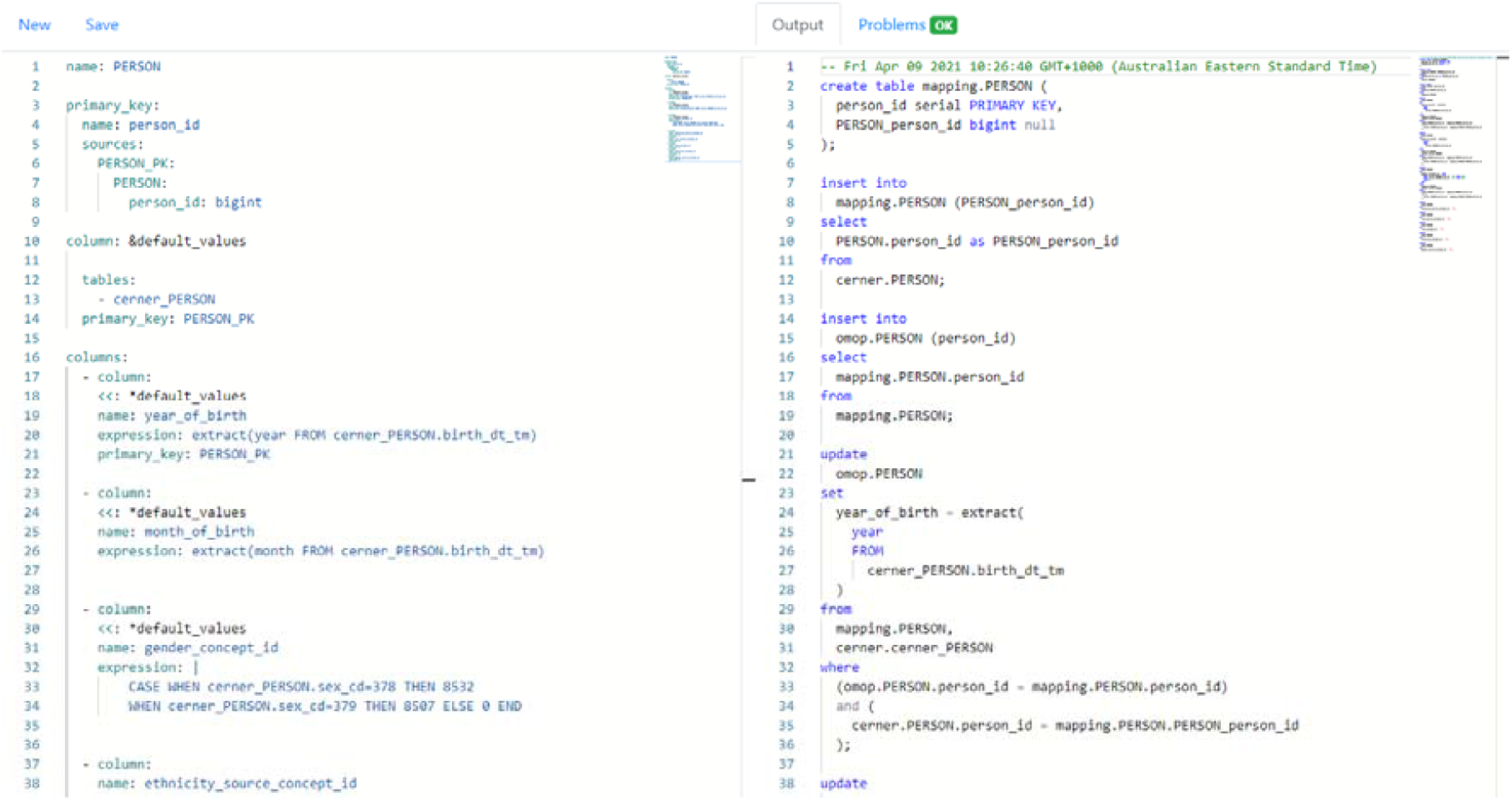
The web application allows users to enter their YAML mapping logic, which gets converted to an ETL SQL script that can be executed in a deployed environment. This is accessible at https://www.omop.link

### Validation Details

**Figure 1.**
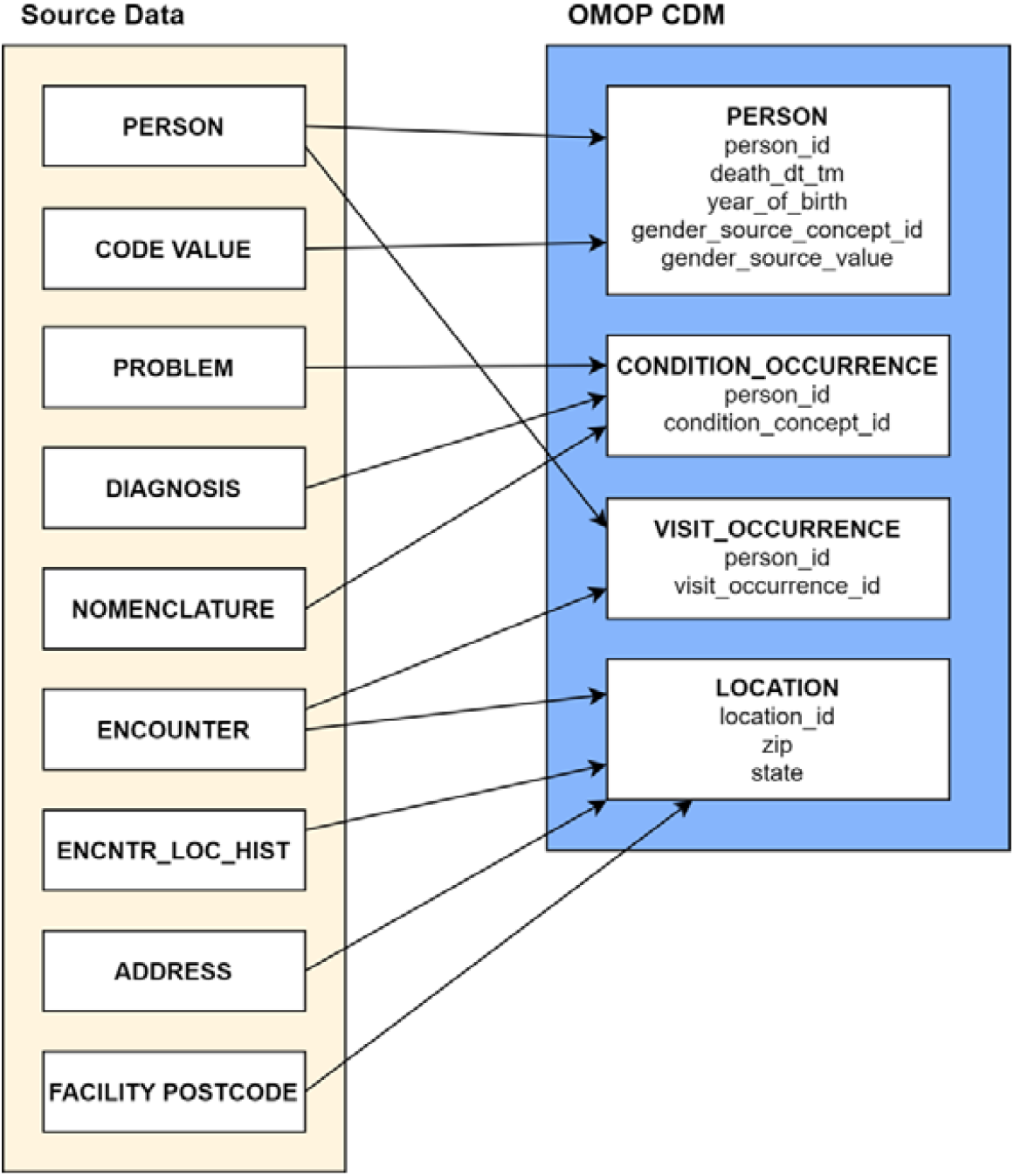
Mapping of simulated CERNER data to OMOP for validating our DML and ETL framework. This mapping captures the data conversion to test all features of the ETL framework.

